# ZIBGLMM: Zero-Inflated Bivariate Generalized Linear Mixed Model for Meta-Analysis with Double-Zero-Event Studies

**DOI:** 10.1101/2024.07.25.24310959

**Authors:** Lu Li, Lifeng Lin, Joseph C. Cappelleri, Haitao Chu, Yong Chen

## Abstract

Double-zero-event studies (DZS) pose a challenge for accurately estimating the overall treatment effect in meta-analysis. Current approaches, such as continuity correction or omission of DZS, are commonly employed, yet these ad hoc methods can yield biased conclusions. Although the standard bivariate generalized linear mixed model can accommodate DZS, it fails to address the potential systemic differences between DZS and other studies. In this paper, we propose a zero-inflated bivariate generalized linear mixed model (ZIBGLMM) to tackle this issue. This two-component finite mixture model includes zero-inflation for a subpopulation with negligible or extremely low risk. We develop both frequentist and Bayesian versions of ZIBGLMM and examine its performance in estimating risk ratios (RRs) against the bivariate generalized linear mixed model and conventional two-stage meta-analysis that excludes DZS. Through extensive simulation studies and real-world meta-analysis case studies, we demonstrate that ZIBGLMM outperforms the bivariate generalized linear mixed model and conventional two-stage metaanalysis that excludes DZS in estimating the true effect size with substantially less bias and comparable coverage probability.

## 1 Introduction

Meta-analysis serves as an important tool for synthesizing evidence from multiple studies, providing a systematic and comprehensive understanding of the overall treatment effect. However, double-zero-event studies (DZS) — those with no events in both arms — present critical statistical challenges, leading to potential numerical instability and bias in estimating treatment effects.^1,2^ Such studies are particularly prevalent in fields associated with rare events, such as surgical complications or adverse drug reactions.^3–5^

Various strategies have been proposed for handling single-zero-event or double-zero-event studies in meta-analysis. For common effect size measures such as risk ratio (RR) and odds ratio (OR), there are divergent views on how to handle DZS. When there are zero events in one arm, the continuity correction with a fixed value of 0.5 is commonly added to each cell of a 2 by 2 table for those studies.^1,6^ This approach allows the calculation of effect sizes such as risk ratios and odds ratios without encountering division by zero. If there are zero events in both arms, conventional practice omits these DZS from meta-analyses of OR and RR, arguing that they contribute no information to the magnitude of the treatment effect.^1^ Bayesian approaches have also been proposed.^7,8^ However, even with non-informative priors, selecting the right prior distribution is crucial as it can significantly impact the analysis results, particularly in cases involving rare events.^9–11^

If risk difference is of interest, Tian et al.^12^ developed an exact inference procedure to synthesize evidence from DZS in a meta-analysis. Rücker et al.^13^ proposed to use the arcsine difference as a way to define treatment effects in meta-analyses with double-zero studies. However, the limitation of the arcsine difference method is that the practical usage and interpretation of this difference in the arcsine scale is restricted.^14^

Notably, while continuity correction and omission are commonly used for handling DZS, these methods can lead to biased conclusions.^13^ Both the statistical significance and the direction of intervention effect can change after excluding DZS, as shown in both simulation studies and empirical data analyses.^14–16^ Though the continuity correction method can avoid computational errors, it usually could bias study estimates towards no difference and over-estimate variances of study estimates.^4^ In addition, using different continuity corrections may result in different conclusions.^1^

Excluding DZS is straightforward to implement but may lead to misleading inference. Böhning and Patarawan showed that double-zero studies do not contribute the conditional log likelihood when using one-stage method using a Poisson regression model and a conditional binomial model to include the double-zero studies.^17^ However, as Xu et al.^3^ pointed out, this may not hold true for some other models, such as the multilevel logistic regression model,^18^ beta-binomial model.^19^ In addition, excluding DZS does not fully utilize all the available evidence and can potentially lead to misleading inference if the excluded studies are systematically different from the included studies.^3,20–22^ Also, If the assumed underlying population event probabilities are not zero, DZS contain information for inference on the parameters such as the common odds ratio in meta-analysis and can contribute to the estimation of treatment effects and thus cannot be left out in our analysis.^23^ Figure 1(A) illustrates how the effect sizes could change significantly depending on whether we include or exclude double zero studies based on 1,111 meta-analyses from Cochrane Database of Systematic Reviews (CDSR); see details in Section 5.

**Figure 1:**
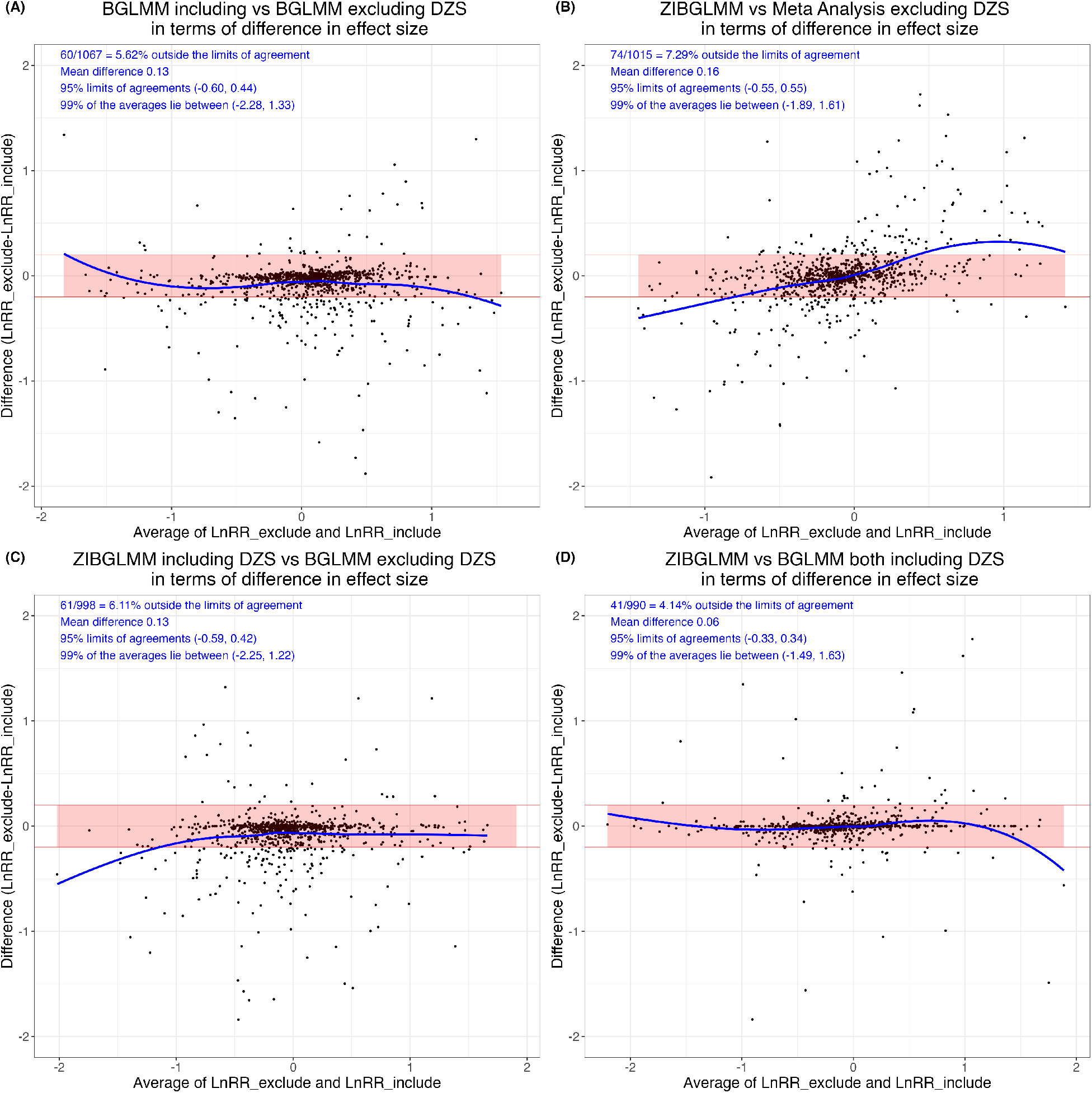
Estimated effect size differences using four methods, i.e., bivariate generalized linear mixed model (BGLMM) including double-zero-event studies (DZS), BGLMM excluding DZS, conventional two-stage meta-analysis excluding DZS (MA), and zero-inflated bivariate generalized linear mixed model (ZIBGLMM) including DZS. In each subfigure, the y-axis is the difference in log RRs between the two methods, and the x-axis is the average of the log RRs of the two methods being compared. Subfigure (A) contrasts the BGLMM with and without DZS. Subfigure (B) explores the difference in effect size between ZIBGLMM and MA, both excluding DZS. Subfigure (C) presents a similar comparison between ZIBGLMM including DZS and BGLMM excluding DZS. Subfigures (A)-(C) shed light on how the inclusion or exclusion of DZS significantly impacts effect sizes based on 1,111 Cochrane meta-analyses. Subfigure (D) provides a comparison between ZIBGLMM and BGLMM, both incorporating DZS. It illustrates a more concentrated distribution, signifying a smaller difference in log RR. Each subfigure displays the number of meta-analyses lying outside the 95% limits of agreement, the mean difference between log RRs, 95% limits of agreements, and 99% range of the averages of log RRs at its upper left corner.

Generalized linear mixed models (GLMM) offer a flexible approach for modeling effect sizes and can incorporate information from DZS without ad hoc continuity correction.^18,24^ Bivariate generalized linear mixed models (BGLMM) have been proposed to include random effects and potential correlation between treatment groups.^25–28^ These models can handle studies with zero events by specifying an appropriate link function and error distribution. For example, BGLMM has been applied to assess whether including DZS impacts the conclusions in a recent systematic review on prevention measures for preventing person-to-person transmission of COVID-19.^29^

Despite their utility, all the models mentioned above fail to address one potential key cause of DZS, that is, non-exchangeable heterogeneity in the population. DZS may occur if the study involves subpopulations with a negligible or extremely low probability of experiencing the event of interest. For instance, healthy subjects less than 65 years old only have negligible risks of experiencing hospitalization or death due to severe symptoms from COVID-19, compared to immunocompromised, unhealthy, or older subjects. The study populations with negligible or extremely low risks are fundamentally different from the remaining populations with low or moderate risks due to intrinsic differences in patient characteristics. Since the BGLMM assumes study populations are exchangeable, the above heterogeneity cannot be modeled properly with BGLMM.

To appropriately handle DZS and account for non-exchangeable heterogeneity in study populations, we propose a zero-inflated bivariate generalized linear mixed model (ZIBGLMM). Zero-inflated models have been commonly applied in other areas to model excess zero counts. Zero-inflated Poisson (ZIP) models have been applied to random-effects meta-analysis.^30^ Beisemann et al.^31^ compared three models (random-effects (RE) Poisson regression, RE zero-inflated Poisson regression, binomial regression) to the standard methods in conjunction with different continuity corrections and different versions of beta-binomial regression.

Our paper is the first to apply zero-inflated models in meta-analyses to address non-exchangeable heterogeneity across study populations. It assumes that a meta-analysis with a proportion of zero-event studies potentially contains two subpopulations: one with a near-zero risk and another with a higher risk. The ZIBGLMM can account for non-exchangeable heterogeneity as well as the correlation among studies in a data-driven fashion. Further, it can properly estimate the overall effect size by avoiding the biases in ad hoc solutions, and can accurately infer the proportion of the low-risk population in each meta-analysis study.

Our contributions in this paper are twofold. First, we introduce a novel method, ZIBGLMM, to incorporate DZS into meta-analyses. Our model takes into account potential population heterogeneity and demonstrates its utility through both real-world and simulation studies. Second, we provide both frequentist and Bayesian implementations of the model. SAS and R implementations are publicly available in the GitHub repository.^32^

The rest of the article is organized as follows. Section 2 provides a motivating example that involves numerous DZS with various sample sizes ranging from 22 to 144 to demonstrate the rationale behind ZIBGLMM. Section 3 introduces the zero-inflated models and formulates both a frequentist and a Bayesian version of the model. Section 4 reanalyzes the example case study in Section 2 to demonstrate the clinical usefulness of the proposed method. Sections 5 and 6 present 1,111 real-world meta-analyses from CDSR and 18,000 simulated meta-analyses to compare the performance of various methods for estimating the risk ratios under various scenarios. Finally, Section 7 summarizes our key findings and limitations.

## 2 A Motivating Example

We explain the rationale behind the ZIBGLMM method using an example meta-analysis taken from the CDSR. The study investigates whether misoprostol could help prevent or treat excessive bleeding and reduce maternal deaths among women after birth.^33^ This review involves 19 studies, 5 of which are DZS. These DZS, with sample sizes ranging from 100 to 900, were excluded from the meta-analysis. Table 1 displays the specific data for these studies. The original analysis suggests that when comparing misoprostol using 600 g misoprostol or more versus placebo or other uterotonics, the results for ‘maternal death or severe morbidity’ is statistically non-significant (RR 1.67, 95% CI 0.80 to 3.45). In Section 4, we will revisit this case study to illustrate how including these DZS could yield different clinical conclusions.

**Table 1:** Motivation example study data.^33^.

| Study ID | Experimental<br>Number of Events | Experimental<br>Sample Size | Control<br>Sample Size | Control<br>Number of Events | Posterior Probability of<br>Belonging to the Structural-Zero |
| --- | --- | --- | --- | --- | --- |
| 1 | 10 | 407 | 4 | 402 | 0 |
| 2 | 0 | 1026 | 1 | 1032 | 0 |
| 3 | 1 | 30 | 0 | 30 | 0 |
| 4 | 66 | 488 | 0 | 490 | 0 |
| 5 | 0 | 257 | 0 | 257 | 0.98 |
| 6 | 0 | 79 | 2 | 81 | 0 |
| 7 | 4 | 630 | 3 | 599 | 0 |
| 8 | 1 | 330 | 0 | 331 | 0 |
| 9 | 1 | 97 | 5 | 99 | 0 |
| 10 | 0 | 100 | 0 | 100 | 0.73 |
| 11 | 2 | 812 | 2 | 808 | 0 |
| 12 | 0 | 900 | 0 | 900 | 1 |
| 13 | 1 | 29 | 0 | 32 | 0 |
| 14 | 0 | 533 | 0 | 583 | 1 |
| 15 | 1 | 32 | 1 | 32 | 0 |
| 16 | 5 | 117 | 0 | 121 | 0 |
| 17 | 28 | 705 | 13 | 717 | 0 |
| 18 | 0 | 702 | 0 | 698 | 1 |
| 19 | 15 | 9225 | 15 | 9230 | 0 |

The rationale of the ZIBGLMM method is based on the observation that in a meta-analysis with a low overall event probability, the probability of encountering large studies with double-zero events should be low. For instance, given an event probability of 1%, the likelihood of a DZS with a sample size of 1,000 is approximately 0.99^1000^≈ 4*/*100, 000. Thus, it is improbable that these large DZS belong to the same population as the other studies included in the meta-analysis. This suggests that using the BGLMM method to incorporate all DZS and treating them as exchangeable with other studies may be inappropriate. Conversely, some smaller DZS may have double-zero events by chance, which means that completely excluding all DZS may not be the best approach either. The ZIBGLMM method addresses these issues by using a data-driven approach to identify the proportion of populations with extremely low risks and model population heterogeneity.

The proposed method is only intended to be used for meta-analyses with a moderate to large size (≥10 studies). We acknowledge that most of the real-world meta-analyses are of smaller sizes.^34,35^ However, the sophistication of the proposed statistical model, which includes a bivariate outcome, a 2×2 matrix of random effects, and a mixture distribution, might lead to convergence issues in more realistic scenarios with fewer studies. Although this will potentially limit the broad impact of our method, we would like to avoid misleading conclusions when applying sophisticated models like ours to a limited number of studies.

## 3 Methods

### 3.1 Notation

Let *N*_*ik*_ be the number of subjects, and *P*_*ik*_ be the probability of an event for the *i*^th^ study (*i* = 1, 2, …, *m*) where *k* = 1 represents the treatment (or exposed) group and *k* = 0 represents the control (or unexposed) group, respectively. Let *X*_*ijk*_ denote a Bernoulli random variable with a value of 1 denoting an event and a value of 0 denoting a non-event for the *j*^th^ subject (*j* = 1, 2, …, *N*_*ik*_) of the *i*^th^ study in the *k*^th^ treatment group. Let 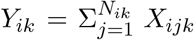 be the total number of events in the *k*^th^ treatment group in the *i*^th^ study. The event counts *Y*_*ik*_ follow a binomial distribution, *Y*_*ik*_ ∼ *Bin* (*N*_*ik*_, *P*_*ik*_). Denote *n* as the total number of studies within a meta-analysis. The notations are summarized in Table 2.

**Table 2:** Descriptions of notation.

| Notation | Description |
| --- | --- |
| $i$ | Index of study |
| $\pi$ | proportion of zero-inflation |
| $k$ | Index of treatment arm, $k = 0$ represents control group and $k = 1$ represents treatment group |
| $Y_{ik}$ | No. of events in the $k^{\text{th}}$ group in the $i^{\text{th}}$ study |
| $n$ | No. of studies in a meta-analysis |
| $N_{ik}$ | No. of subjects in the the $k^{\text{th}}$ group in the $i^{\text{th}}$ study |
| $P_{ik}$ | Probability of an event for the $k^{\text{th}}$ group in the $i^{\text{th}}$ study |
| $\mu_k$ | Fixed effects on the logit scale for the $k^{\text{th}}$ group across entire population |
| $\nu_{ik}$ | Random effects on the logit scale for the $k^{\text{th}}$ group in the $i^{\text{th}}$ study |

### 3.2 Zero-inflated models

The zero-inflated model proposed by Lambert (1992)^36^ has been widely applied to count data with excess zeros in various scientific fields such as industrial manufacturing,^36^ biomedical horticulture,^37^ and healthcare data.^38^ It is formulated as a mixture of a point mass at zero, which generates the excess zeros, and a count distribution that generates the remaining values. The zero-inflated models are particularly useful for data with a higher-than-expected number of zeros under standard count models. Besides commonly used zero-inflated Poisson models, Ghosh et al.^39^ extend the models to include a broad class of distributions (e.g. power series distributions). Böhning et al.^30^ apply the zero-inflated Poisson to random-effects meta-analysis. To the best of our knowledge, this work is the first application of zero-inflated models for handling DZS in the context of meta-analyses with two arms using BGLMM.

Zero-inflated models, by design, segregate observed zeros into two distinct categories. The first category, often referred to as “structural” zeros, represents individuals who are not susceptible to a specific event, thereby having no chance of a positive count. The second category, known as “at-risk” or “chance” zeros, corresponds to a latent group of individuals who are at risk for an event but have a recorded count of zero. The zero-inflated binomial model can then be formulated as follows:

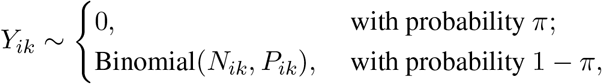

where *π* is the proportion of zero-inflation, i.e., the probability that a given subject belongs to the low-risk subpopulation.

For instance, in our study examining the number of people experiencing adverse events for probiotics, structural zeros might be indicative of patients in good health, resulting in experiencing no adverse events. Conversely, the at-risk zeros could represent patients who are more susceptible to adverse events, due to various circumstances and experienced no adverse events by chance. Consequently, zero-inflated models can be interpreted as latent class models, where the classes are defined by these two types of zeros.^40^

Hall (2020)^37^ discusses classical statistical approaches that utilize the maximum likelihood estimation (MLE) and the likelihood ratio (LR) test for zero-inflated Poisson regression. In the context of non-normal data, classical inference often relies on approximation theory, which is based on large sample sizes and may involve the application of nonstandard asymptotic theory.^41^ However, Ghosh et al.^39^ showed that the classical procedure does not perform as well in estimating the zero-inflation probability when the sample size is finite, and the zero-inflation probability is close to unity, but the Bayesian estimates performed very well with respect to interval width and coverage probability.

### 3.3 Bivariate generalized linear mixed models

To estimate the effect sizes using conventional two-stage meta-analysis, one needs to estimate the event probabilities as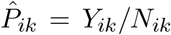. For DZS, 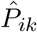 are zero for both the treatment and control arms, creating numerical difficulty in estimating the effect sizes using risk ratios or odds ratios due to division by zero.

One approach to include DZS is to use the BGLMM method to directly model the event counts *Y*_*ik*_ with binomial likelihoods instead of estimating the effect sizes of individual studies. The BGLMM can be specified as follows: let *g* (·) denote the link function that transforms event probabilities into linear forms. We have

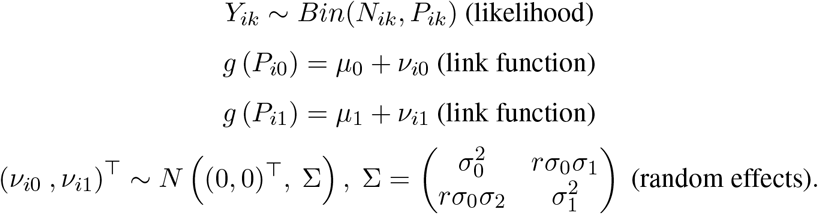

To implement the natural constraint of −1 *< r <* 1, one can use Fisher’s *z* transformation as 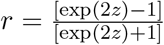.

We consider the logit link function for *g* (·). The parameters *μ*_0_ and *μ*_1_ are fixed effects and represent the average risks in the control and treatment groups in the logit scale. Study-specific random effects in the logit scale, *v*_*i*0_ and *v*_*i*1_, are assumed to follow a bivariate normal distribution with a covariance matrix Σ. The parameters 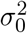 and 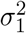 are between-study variances for the control and treatment groups due to heterogeneity, respectively, and *r* is the correlation between the two groups.

### 3.4 Zero-inflated bivariate generalized linear mixed models

One important limitation of the above BGLMM is its inability to account for excess DZS. This is because BGLMM ignores the intrinsic difference between DZS versus the others by treating all studies as exchangeable, even if some studies may be conducted on different subpopulations.

To take into account the population heterogeneity, we introduce the ZIBGLMM method, which is a two-component finite mixture model. Specifically, we assume that the event probability in a certain study subpopulation, referred to as a “healthy population", is extremely low, approximately equal to zero. In contrast, we assume the other subpopulation, referred to as a “sicker population", has a relatively high event probability. We denote *π* as the proportion of studies with healthy populations representing individuals who have approximately zero risk for the event of interest.

In the first stage, the ZIBGLMM combines two zero-generating processes for the number of events *Y*_*ik*_. The first process generates double zeros for both arms from extremely low-risk subpopulations. The second process is governed by a binomial distribution that generates the numbers of events, some of which may be zero by chance. The mixture is described as follows:

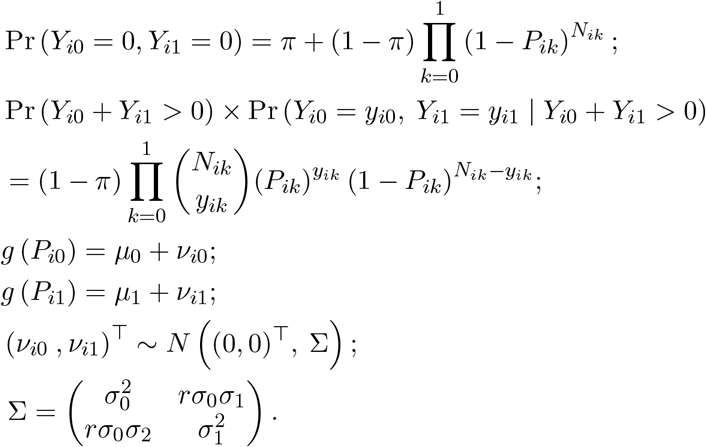

Of note, Dong et al.^42^ proposed a zero-inflated binomial (ZIB) model for meta-analysis with sparse binary outcomes. However, they allowed for different proportions of zero-inflation for different treatment arms, which might not be the case for randomized control trials (RCTs), where the proportion of zero-inflation for the two treatment arms should be the same. In contrast to their approach, we restrict the proportion of the healthy subpopulation to be equal across two treatment arms, which is more realistic given the setting of RCTs.

In this article, we focus on the overall treatment effect in the at-risk population measured by the marginal risk ratio as suggested by McCullagh,^43,44^ which is defined as 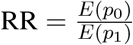. The marginal event probabilities can be obtained through a well-established approximation formula: 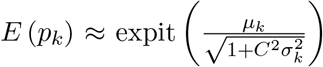 for *k* = 0, 1, with 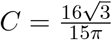 for the bivariate logit random effects model.^45^

Our formulation of ZIBGLMM leverages zero-inflated models to account for subpopulations with an extremely low risk of experiencing the studied outcome. It uses a data-driven approach to capture the heterogeneity in the population, leading to improved model fitting and more reliable results in meta-analyses.

### 3.5 Bayesian formulation

This section introduces a Bayesian formulation of the ZIBGLMM method. It is based on the Bayesian hierarchical model formulation of the random effects meta-analysis model^46,47^ and the Bayesian zero-inflated models.^40^

Specifically, the Bayesian model can be formulated as follows, using the notations in Table 2:

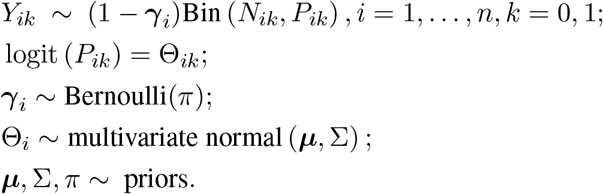

Here, logit(*P*_*i*1_) − logit(*P*_*i*0_) is the log odds ratio. Adopting the same approach, an equivalent model on the risk ratio replaces the logit function with the log function. That is, log(*P*_*i*1_) − log(*P*_*i*0_) is the log risk ratio. We employ a general procedure of data augmentation by including latent variables *γ* to the data *Y* ∼ such that *Y*_*ik*_ = (1 − *γ* _*i*_)*V*_*ik*_ + *γ* _*i*_*W*_*ik*_, where *W*_*ik*_ is the random variable with point mass distribution at 0, *V*_*ik*_ ∼ Bin(*N*_*ik*_, *P*_*ik*_),^48^ i.e., *γ* _*i*_ is a latent variable indicating whether the study is coming from the extremely low-risk population.

The Bayesian approach requires specifying prior distributions for ***µ***, Σ, and *π*. Given the parametrization on logit scale, the event probabilities *P*_*ik*_ are forced to be between 0 and 1, and θ_*ik*_ can range over the whole real line. A natural class of prior distributions for θ_*ik*_ is the class of multivariate normal distributions. We assign the inverse-Wishart distribution, which is a multivariate analog of the gamma distribution, as the semi-conjugate prior distribution for the covariance matrix Σ and the multivariate normal distribution as the prior for ***µ***. We use weakly informative priors for the parameters of ***µ*** and Σ that are weakly centered around estimates derived from the observed data, i.e. the population mean and covariance of *μ*_0_ and *μ*_1_. These are commonly specified in normal hierarchical models.^46^ We use a beta prior for *π*, i.e., *π* ∼ Beta(*a, b*). Note that *a* = *b* = 1 gives the uniform prior on (0, 1) for *π*.

The joint posterior distribution is given by:

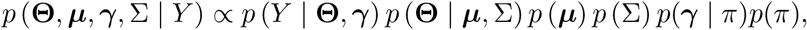

and

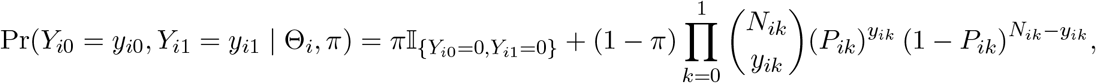

where *P*_*ik*_ = expit(Θ_*ik*_).

We use the Gibbs sampling approach^49^ for the estimation of our full posterior distribution of ***µ***, Σ, *γ, π* by sampling them from their full conditional distributions:

1. *p* (***µ***|Σ, **Θ**, *γ, π, Y*) ∝ *MV N* (***μ***_*n*_, Λ_*n*_), where

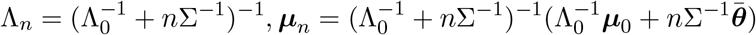

and ***μ***_0_ and Λ_0_ are the prior mean and variance of ***µ***, respectively, and 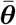 is the vector of treatment-arm-specific averages 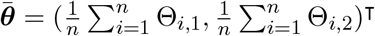.
2. *p*(Σ|***µ*, Θ**, *γ, π, Y*) ∝ inverse-Wishart (η_0_ + *n*, [*S*_0_ + *Sµ*]^−1^), where 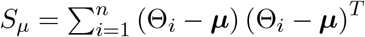, we take *S*_0_ = Λ_0_ but only loosely center Σ around this value by taking η_0_ = *p* + 2 = 4, where *p* is the number of treatment arms.
3. *p*(*γ* _*i*_|**Θ**,***µ***, *π*, Σ, *Y*_*ik*_) ∝ *Bernoulli* 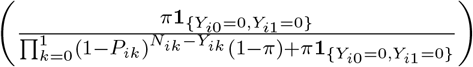 where *P*_*ik*_ = expit(Θ_*ik*_).
4. *p*(*π*|*γ*, **Θ**,***µ***,Σ, *Y*) ∝ *Beta* 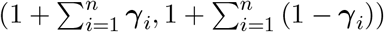.

We then use a Metropolis step^50^ for the estimation of the full posterior distribution of random effects **Θ**.^46,47^ The complete Metropolis-Hastings approximation algorithm is:

1. Sample ***µ***^(*s*+1)^ from *p*(***µ***|Σ^(*s*)^, Θ^(*s*)^, *γ* ^(*s*)^, *π*^(*s*)^, *Y*).
2. Sample Σ^(*s*+1)^ from *p*(Σ|***µ***^(*s*)^, Θ^(*s*)^, *γ* ^(*s*)^, *π*^(*s*)^, *Y*).
3. Sample 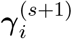 from 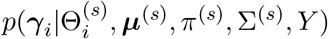 for *i* = 1 … *n*.
4. Sample *π*^(*s*+1)^ from *p*(*π*|*γ* ^(*s*)^, Θ^(*s*)^, ***µ***^(*s*)^, Σ^(*s*)^, *Y*).
5. For each *j* ∈ 1, …, *n*,
  a. sample 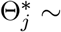 multivariate normal 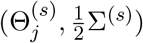 ;
  b. compute the acceptance rate

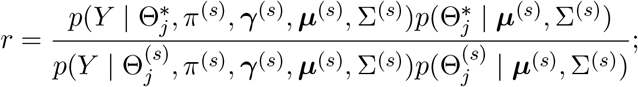
  c. sample *u* ∼ uniform(0, 1). 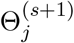 to 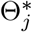 if *u < r* and to 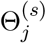 if *u > r*.

We used 100,000 iterations with thinning to keep every 100^th^ value and set the burn-in period to be 10, 000. We manually examined the trace plot and autocorrelation for a few example datasets to make sure the autocorrelation between the samples was low and the acceptance rate was around 50%.

### 3.6 Implementation

The frequentist BGLMM and ZIBGLMM were implemented using PROC NLMIXED in SAS Studio 3.81 (Enterprise Edition). The random effects were approximately integrated by the default adaptive Gaussian quadrature, and likelihood maximization used the conjugate gradient optimization algorithm. The Bayesian models were implemented using R version 4.2.2. In addition to our custom Bayesian Gibbs Sampler, we have implemented a version of the model using the RStan package. This alternative provides a flexible framework, facilitating the customization and experimentation with various prior distributions. The source SAS and R implementation code along with simulation and Cochrane datasets can be found in the GitHub repository.^32^

## 4. Case Study

We apply our ZIBGLMM method to the meta-analysis of 19 studies conducted by Hofmeyr et al.,^33^ which involves 33,041 participants. This meta-analysis compares misoprostol use of greater than or equal to 600 *μg* dose versus placebo in terms of reducing maternal deaths or severe morbidity.^33^ The original analysis suggests that when comparing misoprostol use of greater than or equal to 600 *μg* versus placebo, the results for ‘maternal death or severe morbidity’ are statistically non-significant (RR 1.67, 95% CI 0.80 to 3.45). In the original analysis, 5 of the 19 studies were DZS, with sample sizes ranging from 100 to 900, and were excluded from the analysis.

To evaluate the impact of using different strategies for handling the DZS on the results, we applied various methods to the same data as in Hofmeyr et al. When using the Mantel-Haenszel estimate^51^ of the risk difference, the estimated risk difference is 0.005 when including the DZS and changes to 0.006 when excluding the DZS. The estimated RR using the Mantel-Haenszel method is 2.84 (95% CI 2.03 to 3.99). Our Bayesian ZIBGLMM method yielded an RR of 2.98 (95% CI 1.97 to 4.62), which suggests a 198% increase in the risk of maternal death or severe morbidity associated with high dosage use of misoprostol compared with placebo. This conclusion is also consistent with the general recommendation of using the lowest effective dose of misoprostol to treat/prevent maternal bleeding after giving birth. Using the Bayesian BGLMM method yielded an RR of 2.81 (95% CI 1.78 to 4.37). This investigation underscores that neglecting DZS can lead to non-negligible differences in the final estimated effects in meta-analyses.

We note the difference between the original analysis and the ZIBGLMM method might be due to the fact that they used conditional effects, while we focused on marginal effects.^52^ Nonetheless, the difference between the BGLMM and ZIBGLMM methods is 0.17, suggesting that accounting for DZS could lead to estimates that differ to a large degree.

The estimated proportion of structural zeros *π* is 0.246. For every single study, the posterior probabilities of belonging to the structural-zero or non-structural-zero study cluster are reported in Table 1. The estimated *ρ* is 0.062, which suggests a slight positive correlation between the treatment and control outcomes within studies. The estimated variances are 0.079 and 0.061 for the treatment and control groups, respectively, which provide insights into the variability of the effect sizes within each group. The AIC of the frequentist version of the ZIBGLMM model is 1756.3 and the AIC of the BGLMM model is 1761.7. The DIC for the Bayesian ZIBGLMM model is 979.02, and the DIC for the Bayesian BGLMM is 973.84.

## 5 Cochrane Meta-Analyses

To evaluate the practical impacts of our ZIBGLMM method on a wide spectrum of meta-analyses across different clinical domains, we conducted a meta-meta-analysis using a large number of datasets in the CDSR collected in our previous work.^53^ We identified 1,111 meta-analyses, each containing between 15 and 50 studies (inclusive), with proportions of double-zero studies ranging from 0.15 to 0.4. Figure 2 shows the PRISMA (Preferred Reporting Items for Systematic Reviews and Meta-Analyses) flowchart detailing how we selected the 1,111 studies.^54^ Note that we do not know the true effect sizes for these real-world studies.

**Figure 2:**
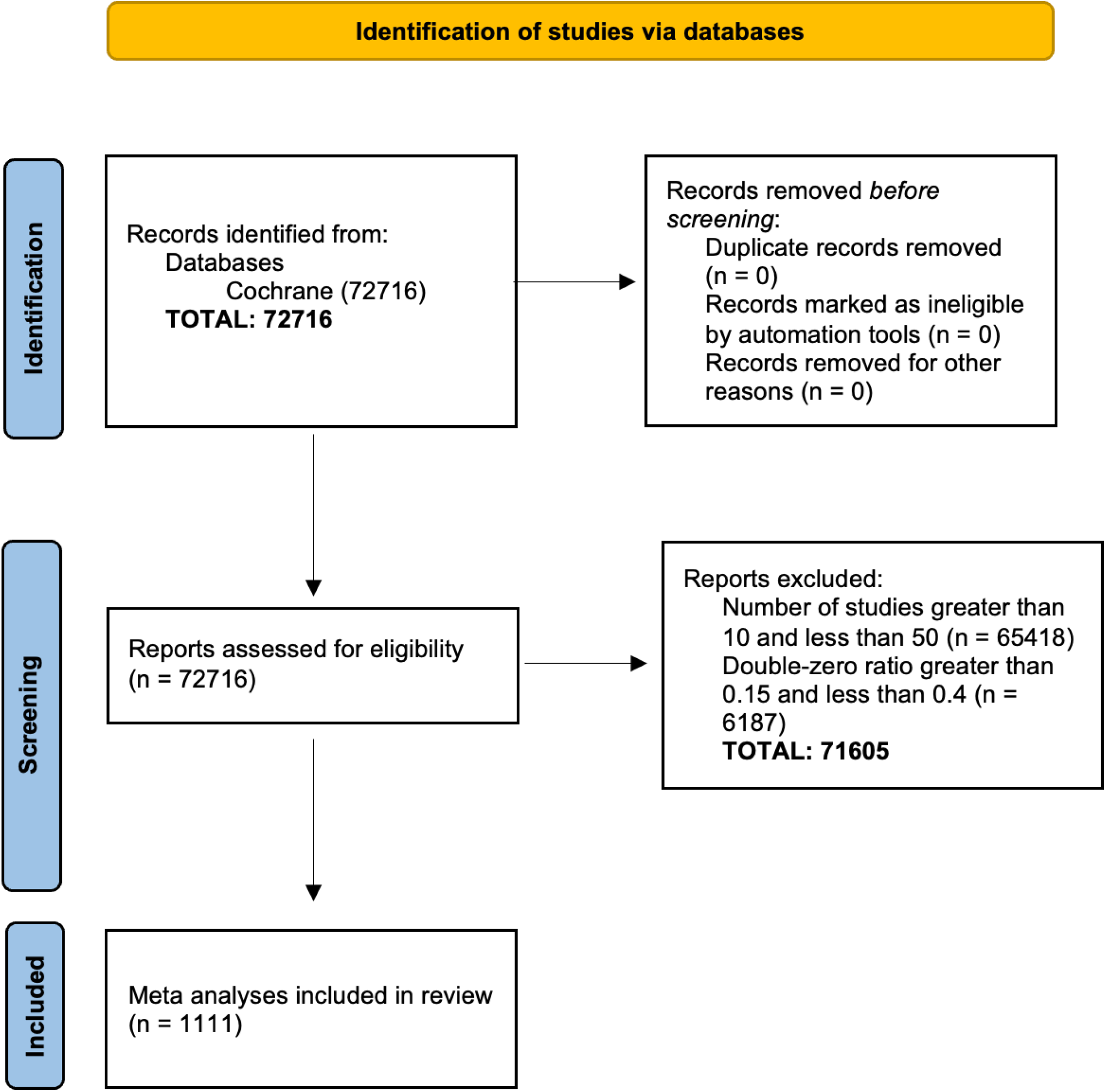
PRISMA plot of the 1,111 Cochrane Database of Systematic Reviews (CDSR) meta-analyses included in this paper. We extracted 1,111 studies with sample sizes between 10 and 50 and with double-zero ratios between 0.15 and 0.4 from a total of 72,716 studies.

We meta-analyzed each dataset using a conventional two-stage method that excludes DZS, as well as both the frequentist and Bayesian versions of BGLMM and ZIBGLMM. When using the frequentist approach, 1,015 datasets converged for ZIBGLMM, while 1,084 datasets converged for BGLMM. The Bayesian BGLMM and ZIBGLMM successfully converged and provided the estimated effect sizes for all 1,111 studies.

To explore the impact of excluding DZS vs. including DZS in a meta-analysis, we contrasted four methods based on the inclusion or exclusion of DZS, focusing on estimated effect size differences. The comparisons are listed below:

A. BGLMM with DZS vs. BGLMM without DZS: to investigate the impact of excluding DZS in one-stage method BGLMM.
B. ZIBGLMM with DZS vs. conventional two-stage meta-analysis without DZS: to investigate the impact of excluding DZS in conventional meta-analysis.
C. ZIBGLMM with DZS vs. BGLMM without DZS: to investigate the impact of including DZS vs. not between two one-stage methods.
D. ZIBGLMM vs. BGLMM, both incorporating DZS: to investigate the impact of using different one-stage methods to incorporate DZS.

Specifically, we applied the Bland-Altman analysis^55^ to determine the difference in each comparison. The results are displayed in Figure 1. Notably, the mean effect size difference can reach up to 0.16 when comparing methods that differ in DZS inclusion (ZIBGLMM with DZS vs. meta-analysis without DZS). However, when both ZIBGLMM and BGLMM include DZS, this mean effect size difference narrowed down to 0.06. This suggests that omitting DZS can significantly alter effect size estimates.

Table 3 presents the results for further quantifying the impact of the exclusion of DZS and the methodological differences in both the magnitude and direction of log RRs. We detail the percentages of increase and decrease and document the count and proportion of pairs with different directions. The effect changed direction if the RRs flipped, this is, log RR changing from “< 0” to “> 0". The significance is flipped if the *p*-value transitioned from nonsignificant to significant, using a 0.05 threshold. We used the middle 99% of the RRs to leave out outliers.

**Table 3:**
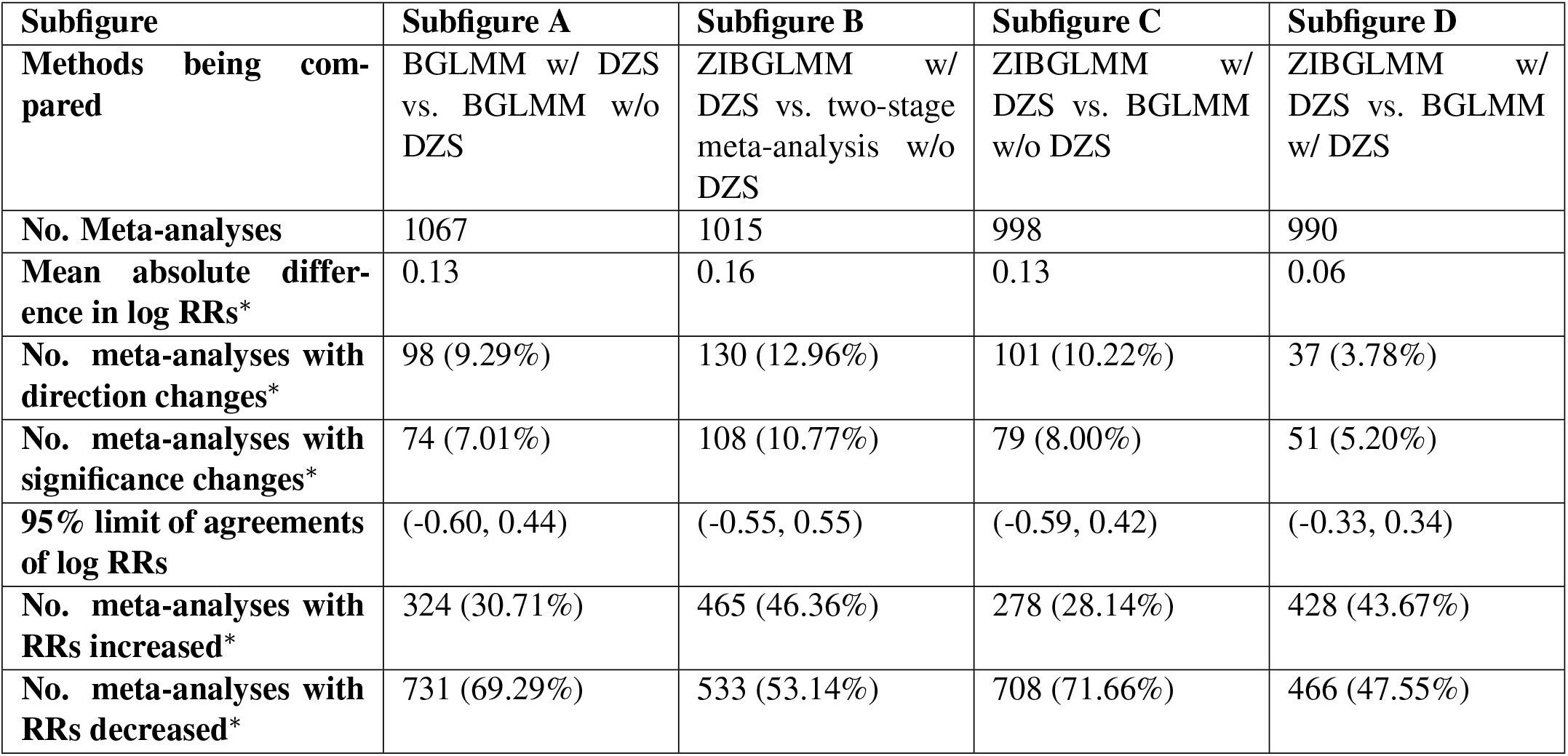
Summary of the comparative analysis of different methods in Figure 1 (^∗^ denotes using the middle 99% of data to exclude extreme values).

The highest percentage of direction change is observed in comparison B (ZIBGLMM with DZS vs. meta-analysis without DZS) at 12.96%, while the lowest percentage is observed in comparison D (ZIBGLMM vs. BGLMM both including DZS) at 3.78%. This indicates excluding DZS can lead to changes in effect directions. Comparison B (ZIBGLMM with DZS vs. meta-analysis without DZS) records the highest percentage of significance flip at 10.77%, with comparison D (ZIBGLMM vs. BGLMM both including DZS) having the lowest at 5.20%. The findings suggest that excluding DZS could lead to significant changes, i.e., difference with a magnitude of greater than 0.1, in estimated effect sizes in meta-analysis. This finding is consistent across various methods, including conventional two-stage meta-analysis that excludes DZS, BGLMM, and ZIBGLMM.

Finally, we compared the frequentist and Bayesian methods of BGLMM and ZIBGLMM in terms of their goodness of fit using the Akaike information criterion (AIC) and deviance information criterion (DIC). Figure 3(B) displays the distribution of AIC and DIC differences for the Cochrane meta-analyses, with the y-axis representing the difference in goodness of fit between ZIBGLMM and BGLMM. The violin plot included a box plot where the box limits indicated the range of the central 50% of the data (i.e., the range between the 25th and 75th percentile), and the median value was marked by a central black line, along with a kernel smoothed density plot representing the probability distribution. ZIBGLMM outperformed BGLMM in 365 of 1,010 (36.13%) Cochrane meta-analyses as measured by AIC, and in 986 of 1,111 (88.74%) datasets based on DIC.

**Figure 3:**
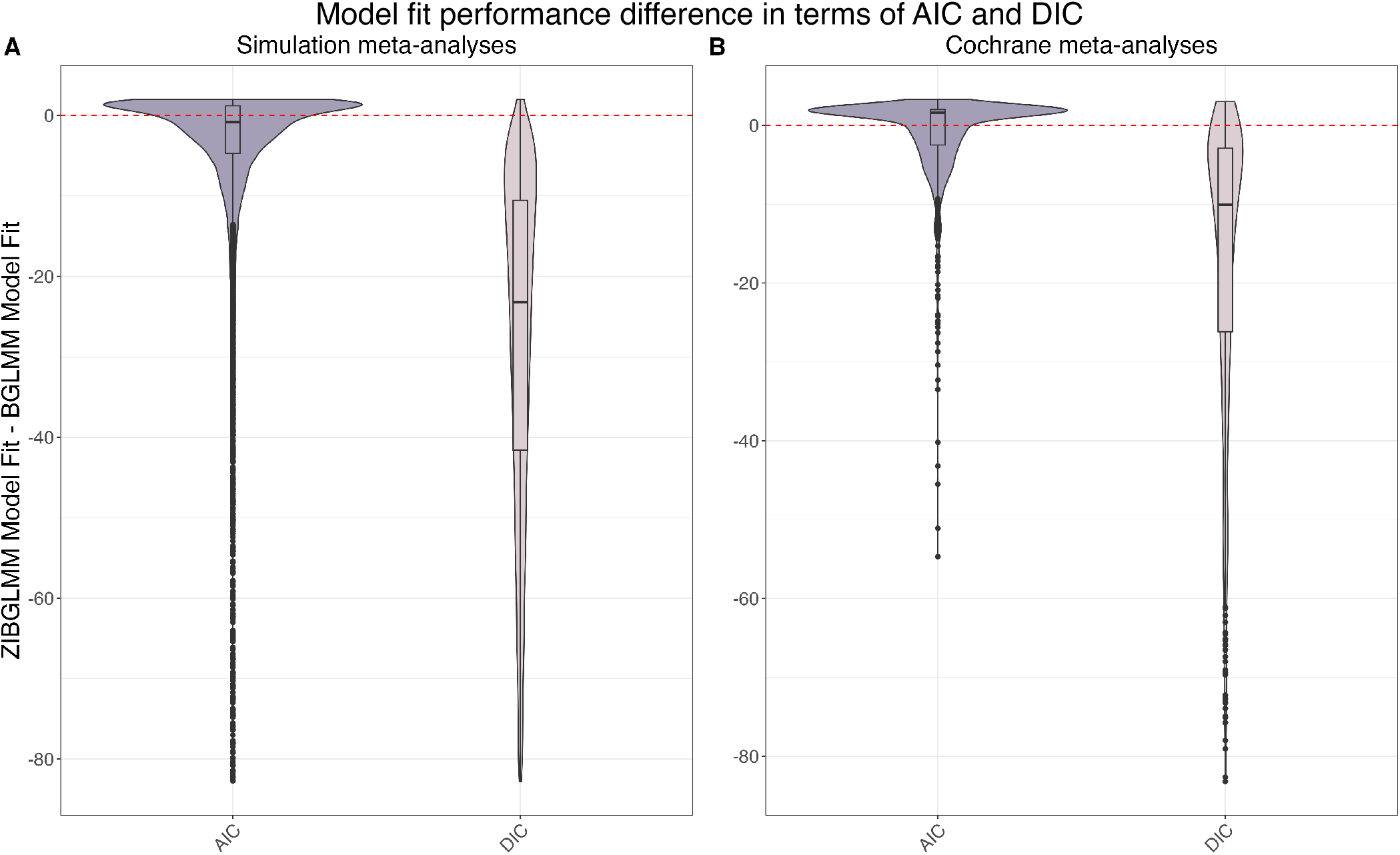
Model goodness of fit in terms of AIC and DIC differences between the ZIBGLMM and BGLMM methods for simulation meta-analyses in subfigure (A) and Cochrane meta-analyses in subfigure (B). Each violin plot^67^ included a box plot where the box limits indicated the range of the central 50% of the data (i.e., the range between the 25th and 75th percentile), and the median value was marked by a central black line, along with a kernel smoothed density plot representing the probability distribution. The y-axis in each subplot represents the goodness of fit of ZIBGLMM subtracting the goodness of fit of BGLMM. The dashed line represents when the difference is 0. Subfigure (A) illustrates the distribution of AIC and DIC differences for 18,000 simulation datasets. The ZIBGLMM exhibited superior fit for 8,839 of 16,215 (54.51%) simulation studies as measured by AIC, and for 17,805 out of 18,000 (98.92%) simulation studies when measured by DIC. Subfigure (B) illustrates the distribution of AIC and DIC differences for 1,111 Cochrane meta-analyses. The ZIBGLMM exhibited superior fit for 365 of 1,010 (36.13%) Cochrane meta-analyses as measured by AIC, and for 986 out of 1,111 (88.74%) Cochrane meta-analyses when measured by DIC.

In summary, our meta-meta analysis study suggests that the inclusion or exclusion of DZS in meta-analyses can influence effect size estimates, with disparities in mean effect size differences reaching up to 0.16. Additionally, both the direction and significance of effect sizes can be altered depending on the methodologies employed. The findings underscore the importance of carefully considering methodological choices in meta-analyses, particularly when handling datasets with double-zero studies.

## 6 Simulations

In addition to the Cochrane datasets, we conducted extensive simulation studies to evaluate our methods in various settings. We simulated meta-analyses with small (10), moderate (25), and large (50) numbers of studies of sample size 50. The non-zero-inflated data were generated using the BGLMM. The number of DZS for each meta-analysis was generated from a binomial distribution with proportions of zero-inflation of 25% and 50%. The baseline event risk in the control group was set at 3% and the average marginal RR was established at 1, 1.5, and 2. Detailed settings of the simulation studies can be found in Table 4. A total of 1,000 meta-analyses were simulated for each of the 18 combinations of settings.

**Table 4:** Specifications for the simulation studies.

| Parameter | Value |
| --- | --- |
| proportion of zero-inflation | 25%, 50% |
| No. of studies | 10 (small), 25 (moderate), 50 (large) |
| Average event probabilities for the non-zero-inflated part | 3% for the control group on average, generated from a BGLMM model |
| Marginal RR | 1, 1.5, 2 |
| Study size | 50 |
| No. of double zero studies | Binomial (# studies, proportion of zero-inflation) |

We applied both the frequentist and Bayesian versions of BGLMM and ZIBGLMM to each simulated meta-analysis, assessing their goodness of fit with AIC and DIC. The distribution of AIC and DIC differences across the 18,000 simulation datasets is depicted in Figure 3(A), with the y-axis representing the goodness of fit differences between ZIBGLMM and BGLMM. The violin plot included a box plot where the box limits indicated the range of the central 50% of the data (i.e., the range between the 25th and 75th percentile), and the median value was marked by a central black line, along with a kernel smoothed density plot representing the probability distribution. ZIBGLMM outperformed BGLMM in 8,839 of 16,215 (54.51%) simulation studies in terms of AIC and in 17,805 of 18,000 (98.92%) studies based on DIC. These findings underscore ZIBGLMM’s robustness in the simulated scenarios.

We compared the coverage properties of conventional two-stage meta-analyses excluding DZS and both the frequentist and Bayesian versions of BGLMM and ZIBGLMM. Figure 4 shows the coverage probabilities, along with the mean lengths of confidence intervals (frequentist models) and credible intervals (Bayesian models), while detailed coverage probabilities can be found in Table 5. Remarkably, the Bayesian BGLMM and Bayesian ZIBGLMM display comparable, consistently high coverage probabilities to meta-analysis, while maintaining the shortest mean confidence interval widths across all settings among all five methods. The coverage probabilities for all methods decrease as the average marginal RRs increase and as the size of studies increases. The coverage properties of frequentist BGLMM and ZIBGLMM were calculated contingent on the number of meta-analyses that attained convergence.

**Table 5:** Coverage probability of the five methods: two-stage meta-analysis excluding DZS (MA), BGLMM, Bayesian BGLMM, ZIBGLMM, and Bayesian ZIBGLMM.

|  |  |  | Frequentist |  | Bayesian |  |
| --- | --- | --- | --- | --- | --- | --- |
| | | | $\pi=0.25$ | $\pi=0.5$ | $\pi=0.25$ | $\pi=0.5$ |
| MA | n=10 | RR=1 | 98.3% | 98.5% | - | - |
|  |  | RR=1.5 | 97.3% | 96.7% | - | - |
|  |  | RR=2 | 96.3% | 96.3% | - | - |
|  | n=25 | RR=1 | 98.1% | 98.1% | - | - |
|  |  | RR=1.5 | 95.2% | 96.1% | - | - |
|  |  | RR=2 | 91.9% | 93.7% | - | - |
|  | n=50 | RR=1 | 97.6% | 97.8% | - | - |
|  |  | RR=1.5 | 92.4% | 94.9% | - | - |
|  |  | RR=2 | 82.8% | 91.3% | - | - |
| BGLMM | n=10 | RR=1 | 97.2% | 96.8% | 98.3% | 98.8% |
|  |  | RR=1.5 | 96.1% | 96.1% | 97.4% | 98.4% |
|  |  | RR=2 | 95.8% | 94.7% | 97.3% | 96.6% |
|  | n=25 | RR=1 | 92.4% | 92.4% | 94.8% | 94.7% |
|  |  | RR=1.5 | 93.5% | 92.4% | 94.3% | 93.6% |
|  |  | RR=2 | 93.1% | 89.3% | 93.6% | 92% |
|  | n=50 | RR=1 | 91.9% | 92.4% | 92.5% | 93.1% |
|  |  | RR=1.5 | 90.8% | 91.5% | 90.7% | 92.2% |
|  |  | RR=2 | 88.5% | 88.9% | 89.6% | 91.6% |
| ZIBGLMM | n=10 | RR=1 | 94% | 94.3% | 98.3% | 98.6% |
|  |  | RR=1.5 | 91.1% | 93.4% | 96.8% | 98% |
|  |  | RR=2 | 90% | 88.3% | 96.9% | 96.3% |
|  | n=25 | RR=1 | 95.4% | 91.8% | 94.9% | 95.2% |
|  |  | RR=1.5 | 91.1% | 89.9% | 94.6% | 93.9% |
|  |  | RR=2 | 87.2% | 85.1% | 93.6% | 92.1% |
|  | n=50 | RR=1 | 92.7% | 91.9% | 92.2% | 93.4% |
|  |  | RR=1.5 | 89.7% | 88.6% | 91.1% | 92.3% |
|  |  | RR=2 | 85.2% | 80% | 90.1% | 91.7% |

**Figure 4:**
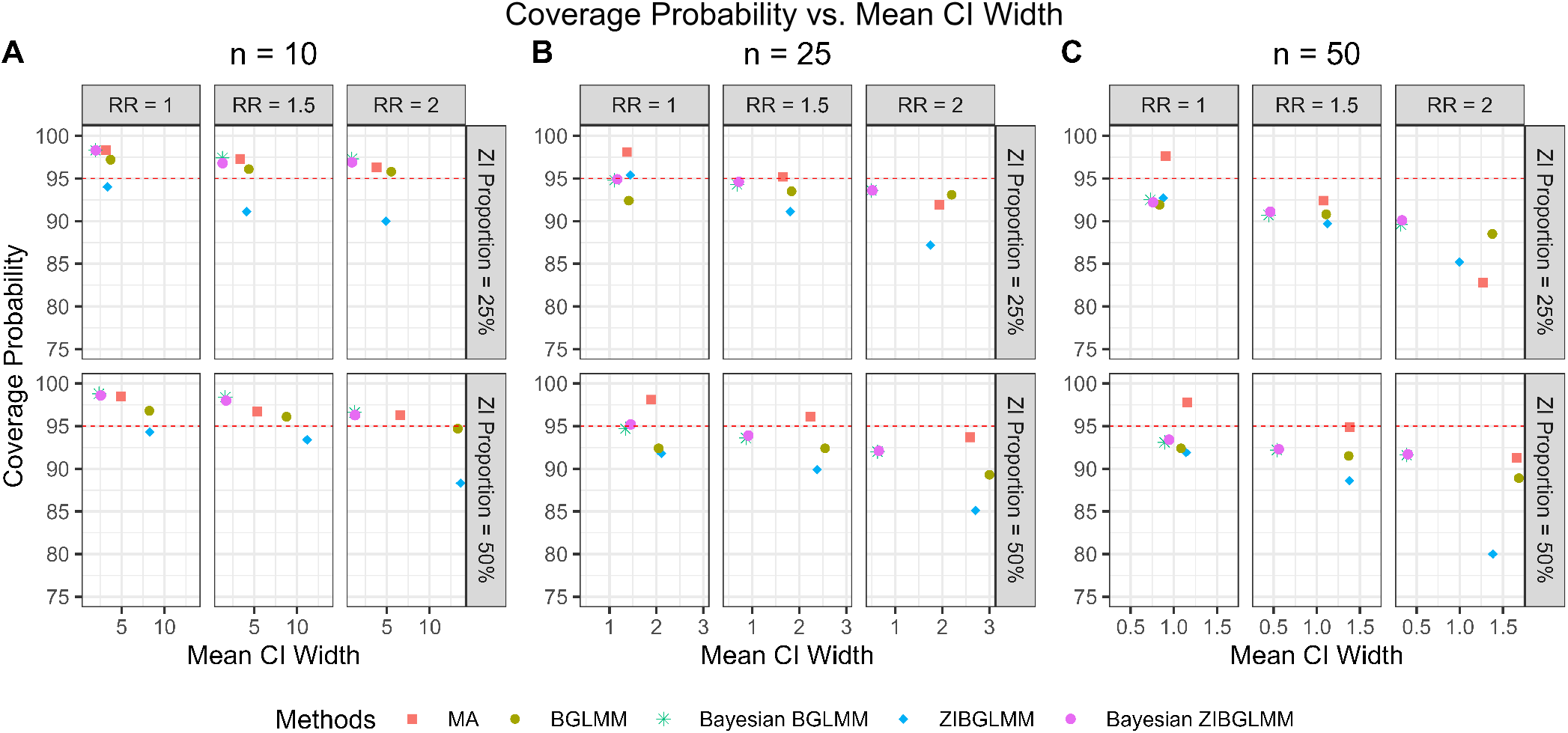
Coverage probability and mean CI length of conventional two-stage meta-analysis (MA), bivariate generalized linear mixed models (BGLMM), Bayesian BGLMM, zero-inflated bivariate generalized linear mixed models (ZIBGLMM), and Bayesian ZIBGLMM. The y-axis displays the coverage probability and the x-axis displays the mean 95% confidence/credible interval widths. The number of studies in a meta-analysis is denoted by *n*. The Bayesian BGLMM and Bayesian ZIBGLMM displayed comparable, consistently high coverage probabilities to MA, while maintaining the shortest mean credible interval widths across all settings. The coverage probabilities for all methods decreased as the average marginal RR increased and as the size of studies increased.

Figure 5 shows the bias in the estimation of RR obtained from conventional two-stage meta-analysis excluding DZS, along with the frequentist and Bayesian versions of BGLMM and ZIBGLMM. The frequentist ZIBGLMM obtains the least bias for meta-analyses of small size (10 studies) while the Bayesian BGLMM manifests the least bias for meta-analyses of moderate size (e.g., with 25 studies) among the five methods. The Bayesian BGLMM and ZIBGLMM obtain effect size estimates with a comparatively small bias for large meta-analyses (e.g., with 50 studies).

**Figure 5:**
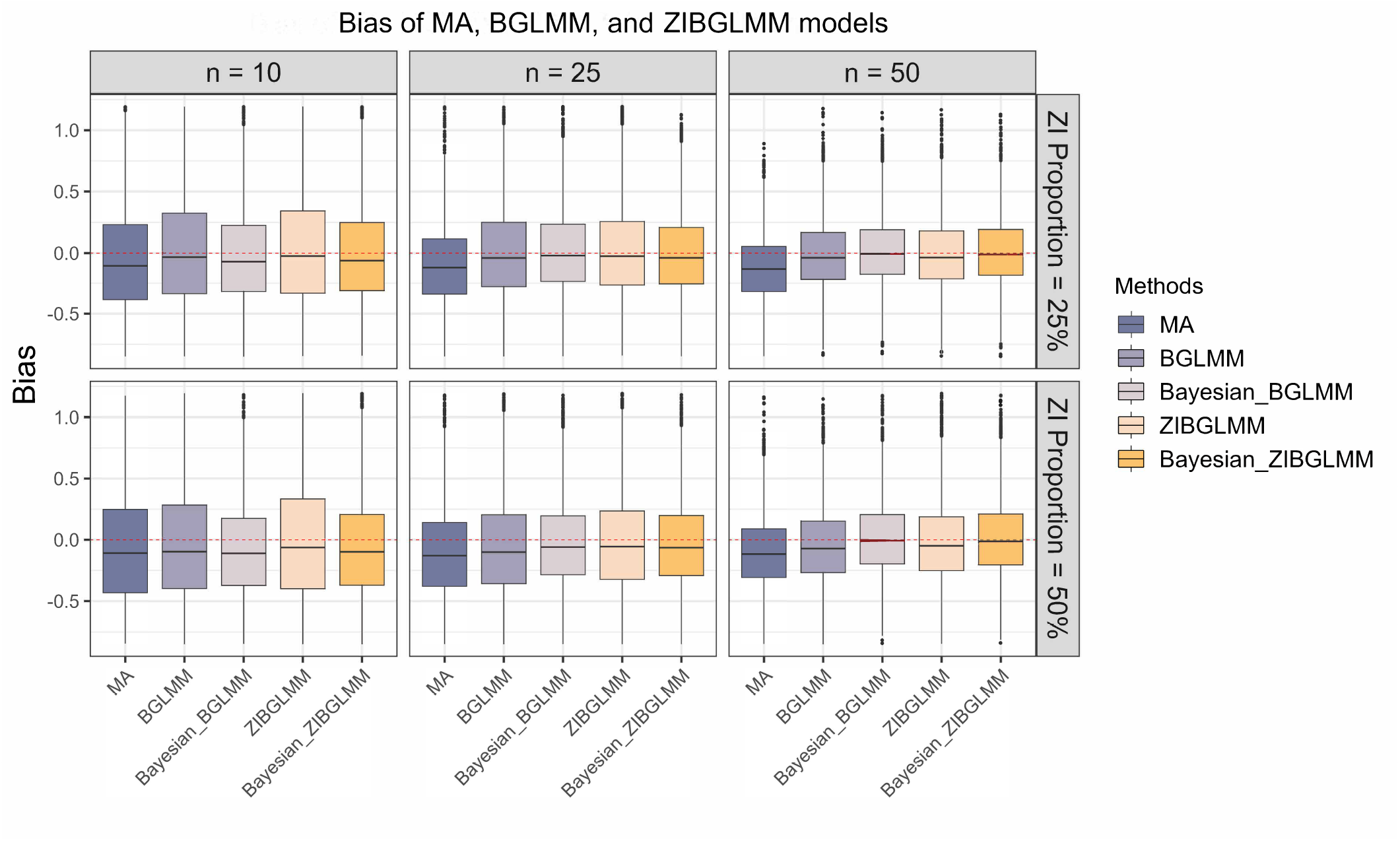
Bias in the estimation of risk ratio (RR) from conventional two-stage meta-analysis excluding DZS (MA), alongside the frequentist and Bayesian versions of BGLMM and ZIBGLMM. For meta-analyses with a smaller size (10 studies), the frequentist ZIBGLMM exhibits the least bias. For meta-analyses with a moderate size (25 studies), Bayesian BGLMM manifests the least bias. The Bayesian BGLMM and ZIBGLMM archives the smallest bias for large (50 studies) meta-analyses. Frequentist ZIBGLMM consistently demon-strates less bias than the frequentist BGLMM. For all settings, both frequentist and Bayesian BGLMM and ZIBGLMM consistently yield smaller biases compared to MA.

Moreover, the frequentist ZIBGLMM consistently demonstrates less bias than the frequentist BGLMM. For all settings, both frequentist and Bayesian BGLMM and ZIBGLMM consistently yielded smaller biases compared to conventional two-stage meta-analyses that exclude DZS. We observe an increased number of convergence issues for frequentist ZIBGLMM, especially for smaller meta-analyses with 10 studies. Possible contributing factors could be the absence of DZS or an excessive presence of DZS within a meta-analysis. Comprehensive details on the convergence issues for *n* = 10 can be found in Table 6. This convergence issue by the zero-inflated models is consistent with the observations made by Beisemann et al.^31^

An important advantage of the proposed ZIBGLMM method is its capability of capturing population heterogeneity by estimating the proportion of zero-inflation. To evaluate the empirical performance of ZIBGLMM under various settings, Figure 6 summarizes the bias of the estimated proportion of zero-inflation with respect to the frequentist and Bayesian ZIBGLMM methods. We observe that Bayesian ZIBGLMM produces estimates of the proportion of zero-inflation with smaller biases than the frequentist ZIBGLMM across meta-analyses of all sizes and proportions of zero-inflation. In addition, Bayesian ZIBGLMM yields more accurate estimates when the proportion of zero-inflation is 50% than when the proportion of zero-inflation is 25%. In contrast, the frequentist ZIBGLMM appears to have a more substantial bias as the size of the meta-analyses grows.

**Figure 6:**
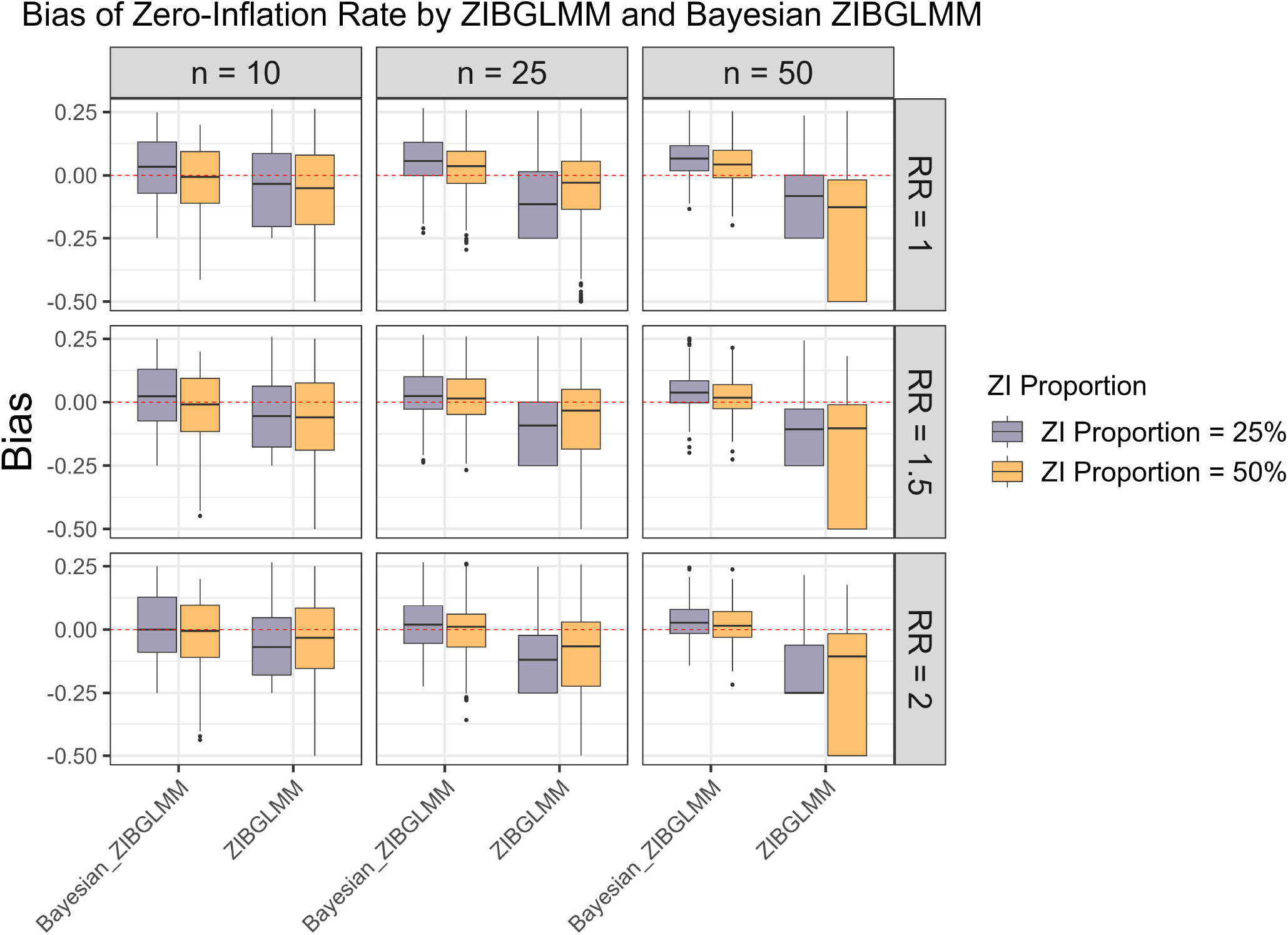
Bias in the estimation of proportion of zero-inflation *π*. Bayesian ZIBGLMM produces an estimate of the proportion of zero-inflation with a smaller bias than the frequentist ZIBGLMM across all study sizes and proportions of zero-inflation. However, it tends to overestimate the proportion of zero-inflation for larger meta-analyses (50 studies). In addition, Bayesian ZIBGLMM yields better estimates when the proportion of zero-inflation is 50% than when it is 25%. The frequentist ZIBGLMM appears to have a more substantial bias as the size of the meta-analyses grows.

In summary, our extensive simulation studies suggest that ZIBGLMM, especially the Bayesian method, is a promising tool for meta-analyses, particularly when dealing with datasets that have varying proportions of DZS. This method provides robust model fit, smaller bias in effect size estimation, and comparatively high coverage probabilities as conventional two-stage meta-analysis. Moreover, the capability of Bayesian ZIBGLMM to estimate the proportion of zero-inflation accurately, especially in smaller and moderate-sized meta-analyses, further emphasizes its significance in capturing the underlying population heterogeneity. However, practitioners need to be cautious while interpreting results from larger meta-analyses, as there may be a tendency to overestimate the proportion of zero-inflation. It’s also worth noting the potential convergence issues faced by the frequentist ZIBGLMM method in certain scenarios, which might be solved by using different optimization algorithms.

## 7 Discussion

In this paper, we proposed a zero-inflated bivariate generalized linear mixed model (ZIBGLMM) as a new method for handling double-zero-event studies (DZS) in meta-analysis. This model is motivated by the hypothesis that zeros in DZS may arise due to heterogeneity in the population, making the segregation of “structural” and “chance” zeros essential for accurate analysis.

The main estimate that we derive from our model is a marginal Relative Risk (RR). Alternatively, one could use the Odds Ratio (OR) as the estimate. We chose RR over OR as the estimate mainly because of the non-collapsibility issues of the OR. There has been an ongoing debate on the choice between OR and RR.^56–59^ Another option is to use a GLMM with a log-link assuming a Poisson distribution within studies as Böhning et al.^60^ to report cluster-specific effects instead of marginal effects.^61^ The choice between reporting the cluster-specific effects versus the marginal effects depends on the population we are interested in generalizing to. Marginal effects should be used if the goal is to generalize to the whole population, while cluster-specific effects should be used if we are only interested in the study-level effects. One potential issue with the bivariate approach to meta-analysis is that it may permit inter-trial information to be recovered, which in theory can lead to bias.^62^ However, as Senn^62^ has pointed out, in practice the size of this bias is likely to be small and can rarely cause an issue in real-world applications.

There are two ways of defining the treatment effects in our model setting. Let *p*_*ik*_ denote the probability of success for the i-th study for the k-th treatment arm. Then, we could estimate the treatment effects based on the overall population (both ZI and non-ZI population), and the formula is *π* + (1 − *π*)*p*_*k*_, where *p*_*k*_ = 𝔼 (*p*_*ik*_). The argument for this approach is that this aligns more with our model of the two mixture parts or only the non-ZI population, and the formula is *p*_*k*_ = 𝔼 (*p*_*ki*_). The argument for this is that since the population is ZI and is healthier, then there are little treatment effects for that population. We have chosen the latter way of defining the treatment effects.

Using 1,111 real-world meta-analyses with DZS selected from the CDSR and 18,000 simulated meta-analyses, we demonstrated the strengths and potential benefits of both the frequentist and Bayesian versions of ZIBGLMM. The Bayesian ZIBGLMM provided consistently better goodness of fit than the Bayesian BGLMM across varying scenarios. We further identified that Bayesian BGLMM and Bayesian ZIBGLMM generally accomplished comparable coverage probabilities to two-stage meta-analyses, shorter confidence interval lengths, and less bias in estimating RR.

The ZIBGLMM method has several limitations compared with the standard bivariate generalized linear model or conventional two-stage meta-analysis excluding DZS. Notably, convergence issues might occur for the frequentist versions of ZIBGLMM and BGLMM models, especially for meta-analyses with a smaller number of studies. This issue potentially stems from the absence of DZS or the excessive presence of DZS in a meta-analysis. The frequentist ZIBGLMM method has a higher chance of encountering convergence issues than the BGLMM method. Specifically, 1,015 datasets converged for ZIBGLMM, while 1,084 datasets converged for BGLMM for the 1,111 CDSR meta-analyses. A potential way of avoiding convergence issues is to change the optimization algorithms used. Alternatively, using the Bayesian BGLMM and ZIBGLMM can avoid the convergence issues.

The limitation of the Bayesian BGLMM and ZIBGLMM is that they typically might take longer time to compute than the frequentist counterparts. For example, fitting a Bayesian ZIBGLMM on a meta-analysis with 25 studies takes around 4 minutes, and fitting on a meta-analysis with 50 studies takes around 9 minutes on a MacBook Pro with an Apple M1 Pro 10-core CPU and 32 GB RAM.

Our general recommendation to practitioners is to opt for Bayesian ZIBGLMM in the presence of double-zero-event studies (DZS), as it provides a more accurate estimation of the risk ratio and the proportion of zero-inflation. Bayesian ZIBGLMM in general achieves better goodness of fit in terms of deviance of information (DIC) than Bayesian BGLMM. In addition, the Bayesian methods are not subject to convergence issues that the frequentist ZIBGLMM might face. The main limitation of the Bayesian methods is that the computation time might be longer for a single meta-analysis than their frequentist counterparts.

If computation time is a major limitation, we recommend using the frequentist ZIBGLMM, which can provide a more accurate estimation of the risk ratio than the frequentist BGLMM. For meta-analyses comprising 10 or fewer studies, we recommend using Bayesian ZIBGLMM for meta-analyses with 10 studies or fewer since the computation time wouldn’t increase too much and the frequentist ZIBGLMM are more likely to experience convergence for the small meta-analyses as observed in the simulation studies. We also recommend exercising caution with large meta-analyses when using Bayesian ZIBGLMM, as there might be a tendency to overestimate the proportion of zero-inflation.

We note that when the true data-generating mechanism (DGM) is BGLMM rather than ZIBGLMM, using ZIBGLMM might be misspecified. In fact, the BGLMM is a submodel of the ZIBGLMM. In practice, if there are excessive numbers of zeros in the data, we recommend the investigators to fit the ZIBGLMM to avoid substantial biases due to underfitting. Further, we can also formally test between the BGLMM and ZIBGLMM models, as they are nested models. Score tests and likelihood ratio tests can be developed following the work by Hall et al.^63^ and Huang et al.^64^ For the likelihood ratio test, one needs to properly account for the fact that under the null hypothesis, the mixing probability parameter *π* = 0 lies at the boundary of its parameter space [0, 1].^41,65,66^

Meta-regression remains a challenging yet vital aspect of enhancing the robustness and applicability of our findings. In particular, when many zero events occur in meta-analyses, the effect sizes tend to be more homogeneous, making it difficult to identify appropriate study-level summary covariates that could explain small between-study heterogeneity. One direction for future work is to expand the current model to incorporate covariate meta-regression, which would allow for a more detailed examination of how specific study characteristics influence the treatment effects.

In conclusion, our study proposes the ZIBGLMM as a novel approach to handling DZS, which can effectively reduce the biases in conventional meta-analysis methods by properly accounting for the between-population heterogeneity. We expect the ZIBGLMM method to be useful in pharmacoepidemiological and pharmacovigilance studies where the event probabilities are rare and DZS are prevalent.

## Data Availability

The data sets and code for this study can be found in the repository:https://github.com/luli2949/ZIBGLMM

## Acknowledgments and Funding

This work was supported in part by National Institutes of Health (R21AI167418, 1R01LM014344, 1R01AG077820, R01LM012607, R01AI130460, R01AG073435, R56AG074604, R01LM013519, R56AG069880, U01TR003709, RF1G077820). This work was supported partially through the Patient-Centered Outcomes Research Institute (PCORI) Project Program Awards (ME-2019C3-18315 and ME-2018C3-14899). All statements in this report, including its findings and conclusions, are solely those of the authors and do not necessarily represent the views of the Patient-Centered Outcomes Research Institute (PCORI), its Board of Governors or Methodology Committee. We thank Tianyu Zhang and Dr. Jiajie Chen for their help on the project.

## Data Availability Statement

The data sets and code for this study can be found in the repository.^32^

## Highlights

What is already known?

- Double-zero-event studies (DZS) are challenging for meta-analysis due to division by zero errors.
- Current methods to address this include continuity correction or omitting DZS.
- These existing methods might produce biased conclusions.

What is new?

- The paper introduces and discusses the rationale for proposing ZIBGLMM (Zero-Inflated Bivariate Generalized Linear Mixed Model), a novel method for handling double-zero-event studies (DZS) in meta-analysis.
- The ZIBGLMM addresses issues by using a data-driven approach to identify subpopulations with extremely low risks and model population heterogeneity.

Potential impact for Research Synthesis Methods readers

- The proposed ZIBGLMM can potentially provide a more accurate estimation of effect sizes, especially in meta-analysis with excess DZS.
- Using a data-driven approach, ZIBGLMM provides a more robust solution to handle DZS compared to existing methods, which could lead to better conclusions in research synthesis.

## Supporting Information

**Table 6:** Summary of the meta-analyses that converged for the frequentist BGLMM and ZIBGLMM methods in the simulation study. *π* denotes the proportion of zero-inflation, *n* is the number of studies in the simulated meta-analysis, and *RR* is the true marginal risk ratio. “# covered” is the number of meta-analyses whose confidence intervals contain the true risk ratio. “# is not NA” is the number of meta-analyses whose random effect and fixed effect for both treatment arms are available using the frequentist methods. “# of converged” is the total number of meta-analyses for which the frequentist methods converged (i.e., the model successfully executes without encountering the “Optimization cannot be completed” error). All the numbers are out of a total of 1,000 simulated meta-analyses.

|  |  |  |  | # covered | # not NA | # of converged |
| --- | --- | --- | --- | --- | --- | --- |
| BGLMM | $\pi = 0.25$ | $n=10$ | $RR=1$ | 847 | 871 | 977 |
| | | | $RR=1.5$ | 819 | 852 | 977 |
| | | | $RR=2$ | 820 | 856 | 984 |
| | | $n=25$ | $RR=1$ | 727 | 787 | 952 |
| | | | $RR=1.5$ | 809 | 865 | 979 |
| | | | $RR=2$ | 825 | 886 | 980 |
| | | $n=50$ | $RR=1$ | 834 | 908 | 981 |
| | | | $RR=1.5$ | 853 | 939 | 991 |
| | | | $RR=2$ | 852 | 963 | 999 |
| | $\pi = 0.5$ | $n=10$ | $RR=1$ | 847 | 875 | 981 |
| | | | $RR=1.5$ | 799 | 831 | 970 |
| | | | $RR=2$ | 815 | 861 | 974 |
| | | $n=25$ | $RR=1$ | 746 | 807 | 958 |
| | | | $RR=1.5$ | 789 | 854 | 977 |
| | | | $RR=2$ | 781 | 875 | 983 |
| | | $n=50$ | $RR=1$ | 843 | 912 | 975 |
| | | | $RR=1.5$ | 866 | 946 | 987 |
| | | | $RR=2$ | 849 | 955 | 993 |
| ZIBGLMM | $\pi = 0.25$ | $n=10$ | $RR=1$ | 772 | 821 | 932 |
| | | | $RR=1.5$ | 703 | 772 | 890 |
| | | | $RR=2$ | 668 | 742 | 860 |
| | | $n=25$ | $RR=1$ | 730 | 765 | 953 |
| | | | $RR=1.5$ | 713 | 783 | 917 |
| | | | $RR=2$ | 635 | 728 | 868 |
| | | $n=50$ | $RR=1$ | 728 | 785 | 939 |
| | | | $RR=1.5$ | 777 | 866 | 955 |
| | | | $RR=2$ | 662 | 777 | 874 |
| | $\pi = 0.5$ | $n=10$ | $RR=1$ | 788 | 836 | 920 |
| | | | $RR=1.5$ | 735 | 787 | 881 |
| | | | $RR=2$ | 651 | 737 | 830 |
| | | $n=25$ | $RR=1$ | 738 | 804 | 898 |
| | | | $RR=1.5$ | 683 | 760 | 875 |
| | | | $RR=2$ | 549 | 645 | 783 |
| | | $n=50$ | $RR=1$ | 718 | 781 | 922 |
| | | | $RR=1.5$ | 697 | 787 | 906 |
| | | | $RR=2$ | 517 | 646 | 760 |

## References

1. J. Sweeting M, J. Sutton A, C. Lambert P. What to add to nothing? Use and avoidance of continuity corrections in meta-analysis of sparse data. Statistics in Medicine. 2004;23(9):1351–1351.

2. Bradburn MJ, Deeks JJ, Berlin JA, Russell Localio A. Much ado about nothing: a comparison of the performance of meta-analytical methods with rare events. Statistics in Medicine. 2007;26(1):53–53.

3. Xu C, Zhou X, Zorzela L, et al. Utilization of the evidence from studies with no events in meta-analyses of adverse events: an empirical investigation. BMC Medicine. 2021;19(1):141.

4. Deeks J J Adge. Chapter 10: Analysing data and undertaking meta-analyses. In: Higgins JPT, Thomas J, Chandler J, Cumpston M, Li T, Page MJ, Welch VA (editors). Cochrane Handbook for Systematic Reviews of Interventions version 6.3 (updated February 2022). Available from https://www.training.cochrane.org/handbook: Cochrane 2022.

5. Tong J, Huang J, Du J, Cai Y, Tao C, Chen Y. Identification of rare adverse events with year-varying reporting rates for FLU4 vaccine in VAERS. in AMIA Annual Symposium Proceedings;2018:1544 American Medical Informatics Association 2018.

6. Cox DR. The continuity correction. Biometrika. 1970;57(1):217–217.

7. Warn DE, Thompson S, Spiegelhalter DJ. Bayesian random effects meta-analysis of trials with binary outcomes: methods for the absolute risk difference and relative risk scales. Statistics in Medicine. 2002;21(11):1601–1601.

8. Vázquez F, Moreno E, Negrín M, Martel M. Bayesian robustness in meta-analysis for studies with zero responses. Pharmaceutical Statistics. 2016;15(3):230–230.

9. Stijnen T, Hamza TH, Özdemir P. Random effects meta-analysis of event outcome in the framework of the generalized linear mixed model with applications in sparse data. Statistics in Medicine. 2010;29(29):3046–3046.

10. Lambert PC, Sutton AJ, Burton PR, Abrams KR, Jones DR. How vague is vague? A simulation study of the impact of the use of vague prior distributions in MCMC using WinBUGS. Statistics in Medicine. 2005;24(15):2401–2428.

11. Senn S. Trying to be precise about vagueness. Statistics in Medicine. 2007;26(7):1417–1417.

12. Tian L, Cai T, Pfeffer MA, Piankov N, Cremieux PY, Wei L. Exact and efficient inference procedure for meta-analysis and its application to the analysis of independent 2× 2 tables with all available data but without artificial continuity correction. Biostatistics. 2009;10(2):275–275.

13. Rücker G, Schwarzer G, Carpenter J, Olkin I. Why add anything to nothing? The arcsine difference as a measure of treatment effect in meta-analysis with zero cells. Statistics in Medicine. 2009;28(5):721–721.

14. Xu C, Li L, Lin L, et al. Exclusion of studies with no events in both arms in meta-analysis impacted the conclusions. Journal of Clinical Epidemiology. 2020;123:91–99.

15. Ren Y, Lin L, Lian Q, Zou H, Chu H. Real-world performance of meta-analysis methods for double-zero-event studies with dichotomous outcomes using the Cochrane database of systematic reviews. Journal of General Internal Medicine. 2019;34:960–968.

16. Cheng J, Pullenayegum E, Marshall JK, Iorio A, Thabane L. Impact of including or excluding both-armed zero-event studies on using standard meta-analysis methods for rare event outcome: a simulation study. BMJ Open. 2016;6(8):e010983.

17. Böhning D, Sangnawakij P. The identity of two meta-analytic likelihoods and the ignorability of double-zero studies. Biostatistics. 2020;22(4):890–890.

18. Platt RW, Leroux BG, Breslow N. Generalized linear mixed models for meta-analysis. Statistics in Medicine. 1999;18(6):643–643.

19. Mathes T, Kuss O. Beta-binomial models for meta-analysis with binary outcomes: variations, extensions, and additional insights from econometrics. Research Methods in Medicine & Health Sciences. 2021;2(2):82–82.

20. Xu C, Lin L, Vohra S. Evidence synthesis practice: why we cannot ignore studies with no events?. Journal of General Internal Medicine. 2022;37(14):3744–3744.

21. Friedrich JO, Adhikari NK, Beyene J. Inclusion of zero total event trials in meta-analyses maintains analytic consistency and incorporates all available data. BMC Medical Research Methodology. 2007;7(1):5.

22. Kuss O. Statistical methods for meta-analyses including information from studies without any eventsadd nothing to nothing and succeed nevertheless. Statistics in Medicine. 2015;34(7):1097–1097.

23. Xie Mg, Kolassa J, Liu D, Liu R, Liu S. Does an observed zero-total-event study contain information for inference of odds ratio in meta-analysis?. Statistics and Its Interface. 2018;11(2):327–327.

24. Xu C, Furuya-Kanamori L, Lin L. Synthesis of evidence from zero-events studies: A comparison of one-stage framework methods. Research Synthesis Methods. 2022;13(2):176–176.

25. Chu H, Nie L, Chen Y, Huang Y, Sun W. Bivariate random effects models for meta-analysis of comparative studies with binary outcomes: methods for the absolute risk difference and relative risk. Statistical Methods in Medical Research. 2012;21(6):621–621.

26. Chu H, Cole SR. Bivariate meta-analysis of sensitivity and specificity with sparse data: a generalized linear mixed model approach. Journal of Clinical Epidemiology. 2006;59(12):1331–1331.

27. Guo J, Xiao M, Chu H, Lin L. Meta-analysis methods for risk difference: A comparison of different models. Statistical Methods in Medical Research. 2023;32(1):3–3.

28. Lin L, Chu H. Meta-analysis of proportions using generalized linear mixed models. Epidemiology. 2020;31(5):713–713.

29. Xiao M, Lin L, Hodges JS, Xu C, Chu H. Double-zero-event studies matter: A re-evaluation of physical distancing, face masks, and eye protection for preventing person-to-person transmission of COVID-19 and its policy impact. Journal of Clinical Epidemiology. 2021;133:158–160.

30. Böhning D, Mylona K, Kimber A. Meta-analysis of clinical trials with rare events. Biometrical Journal. 2015;57(4):633–633.

31. Beisemann M, Doebler P, Holling H. Comparison of random-effects meta-analysis models for the relative risk in the case of rare events: A simulation study. Biometrical Journal. 2020;62(7):1597–1597.

32. Li, Lu. Source code, Zero-inflated bivariate generalized linear mixed effects model (ZIBGLMM). https://github.com/luli2949/ZIBGLMM 2023. Accessed June 2023.

33. Hofmeyr GJ, Gülmezoglu AM, Novikova N, Lawrie TA. Postpartum misoprostol for preventing maternal mortality and morbidity. Cochrane Database of Systematic Reviews. 2013(7).

34. Turner RM, Davey J, Clarke MJ, Thompson SG, Higgins JP. Predicting the extent of heterogeneity in meta-analysis, using empirical data from the Cochrane Database of Systematic Reviews. International journal of epidemiology. 2012;41(3):818–818.

35. Vandermeer B, Bialy L, Hooton N, et al. Meta-analyses of safety data: a comparison of exact versus asymptotic methods. Statistical methods in medical research. 2009;18(4):421–421.

36. Lambert D. Zero-inflated Poisson regression, with an application to defects in manufacturing. Technometrics. 1992;34(1):1–1.

37. Hall DB. Zero-inflated Poisson and binomial regression with random effects: a case study. Biometrics. 2000;56(4):1030–1030.

38. Mouatassim Y, Ezzahid EH. Poisson regression and Zero-inflated Poisson regression: application to private health insurance data. European Actuarial Journal. 2012;2(2):187–187.

39. Ghosh SK, Mukhopadhyay P, Lu JCJ. Bayesian analysis of zero-inflated regression models. Journal of Statistical Planning and Inference. 2006;136(4):1360–1360.

40. Neelon B. Bayesian zero-inflated negative binomial regression based on Pólya-Gamma mixtures. Bayesian Analysis. 2019;14(3):829–829.

41. Self SG, Liang KY. Asymptotic properties of maximum likelihood estimators and likelihood ratio tests under nonstandard conditions. Journal of the American Statistical Association. 1987;82(398):605–605.

42. Dong C, Zhao Y, Tiwari R. Meta-Analysis of Clinical Trials With Sparse Binary Outcomes Using Zero-Inflated Binomial (ZIB) Models. Statistics in Biopharmaceutical Research. 2019;11(3):228–228.

43. McCullagh P. Sampling bias and logistic models. Journal of the Royal Statistical Society Series B: Statistical Methodology. 2008;70(4):643–643.

44. Longford N, Diggle P, Nelder J, et al. Sampling bias and logistic models-Discussion. Journal of the Royal Statistical Society Series B-Statistical Methodology. 2008;70.

45. Zeger SL, Liang KY, Albert PS. Models for longitudinal data: a generalized estimating equation approach. Biometrics. 1988;44(4):1049–1049.

46. Hoff PD. A First Course in Bayesian Statistical Methods;580. Springer 2009.

47. Gelman A, Carlin JB, Stern HS, Dunson DB, Vehtari A, Rubin DB. Bayesian Data Analysis. CRC press 2013.

48. Tanner MA, Wong WH. The calculation of posterior distributions by data augmentation. Journal of the American Statistical Association. 1987;82(398):528–528.

49. Geman S, Geman D. Stochastic relaxation, Gibbs distributions, and the Bayesian restoration of images. IEEE Transactions on Pattern Analysis and Machine Intelligence. 1984;PAMI-6(6):721–741.

50. Metropolis N, Rosenbluth AW, Rosenbluth MN, Teller AH, Teller E. Equation of state calculations by fast computing machines. Journal of Chemical Physics. 1953;21(6):1087–1087.

51. Mantel N, Haenszel W. Statistical Aspects of the Analysis of Data From Retrospective Studies of Disease. JNCI: Journal of the National Cancer Institute. 1959;22(4):719–719.

52. Neuhaus JM, Kalbfleisch JD, Hauck WW. A Comparison of Cluster-Specific and Population-Averaged Approaches for Analyzing Correlated Binary Data. International Statistical Review / Revue Internationale de Statistique. 1991;59(1):25–25.

53. Lin L, Chu H, Murad MH, et al. Empirical Comparison of Publication Bias Tests in Meta-Analysis. J Gen Intern Med. 2018;33(8):1260–1260.

54. Moher D, Liberati A, Tetzlaff J, Altman DG, Group* P. Preferred reporting items for systematic reviews and meta-analyses: the PRISMA statement. Annals of internal medicine. 2009;151(4):264–264.

55. Altman DG, Bland JM. Measurement in medicine: the analysis of method comparison studies. Journal of the Royal Statistical Society: Series D (The Statistician). 1983;32(3):307–307.

56. Xiao M, Chen Y, Cole SR, MacLehose RF, Richardson DB, Chu H. Controversy and Debate: Questionable utility of the relative risk in clinical research: Paper 2: Is the Odds Ratio portable in meta-analysis? Time to consider bivariate generalized linear mixed model. Journal of Clinical Epidemiology. 2022;142:280–287.

57. Xiao M, Chu H, Cole S, et al. Odds Ratios are far from “portable”—A call to use realistic models for effect variation in meta-analysis. Journal of Clinical Epidemiology. 2022;142:294.

58. Doi SA, Furuya-Kanamori L, Xu C, Lin L, Chivese T, Thalib L. Controversy and debate: questionable utility of the relative risk in clinical research: paper 1: a call for change to practice. Journal of clinical epidemiology. 2022;142:271–279.

59. Doi SA, Furuya-Kanamori L, Xu C, et al. The Odds Ratio is portable across baseline risk but not the Relative Risk: Time to do away with the log link in binomial regression. Journal of Clinical Epidemiology. 2022;142:288–293.

60. Böhning D, Mylona K, Kimber A. Meta-analysis of clinical trials with rare events. Biometrical Journal. 2015;57(4):633–633.

61. Dias S, Ades AE. Absolute or relative effects? Arm-based synthesis of trial data. Research Synthesis Methods. 2016;7(1):23–23.

62. Senn S. Hans van Houwelingen and the Art of Summing up. Biometrical Journal. 2010;52(1):85–85.

63. Hall DB, Berenhaut KS. Score tests for heterogeneity and overdispersion in zero-inflated Poisson and binomial regression models. Canadian journal of statistics. 2002;30(3):415–415.

64. Huang J, Cai Y, Du J, et al. Monitoring vaccine safety by studying temporal variation of adverse events using vaccine adverse event reporting system. 2021.

65. Chen Y, Ning J, Ning Y, Liang KY, Bandeen-Roche K. On pseudolikelihood inference for semiparametric models with boundary problems. Biometrika. 2017;104(1):165–165.

66. Chen Y, Liang KY. On the asymptotic behaviour of the pseudolikelihood ratio test statistic with boundary problems. Biometrika. 2010;97(3):603–603.

67. Hintze JL, Nelson RD. Violin Plots: A Box Plot-Density Trace Synergism. The American Statistician. 1998;52(2):181–181.

